# Factors influencing the trustworthiness of non-randomized studies of interventions: a survey of international experts

**DOI:** 10.1101/2025.09.24.25336459

**Authors:** Sally Yaacoub, John P. A. Ioannidis, Asbjørn Hróbjartsson, Raphaël Porcher, Philippe Ravaud, Isabelle Boutron

**Author notes:** **Corresponding author:** Sally Yaacoub, BS Pharm, MPH, PhD Cochrane France, Center for Research in Epidemiology and Statistics (CRESS), Hôpital Hôtel Dieu, 1 place du Parvis Notre-Dame, 75004 Paris, France.

## Abstract

**Background:** Perceived trustworthiness of research may be influenced by factors beyond the risk of bias, including study-related characteristics, research context, and external circumstances. Identifying these factors is essential for gauging the credibility of non-randomized studies of interventions (NRSIs) as they are interpreted and used in systematic reviews, and for improving their design to ensure that they provide reliable evidence for decision-making. Our objective was to identify factors, not covered in risk of bias assessment tools, that could influence the trustworthiness of NRSIs.

**Methods:** We conducted a cross-sectional survey of international experts. We defined trustworthiness as the proper, justified or rational trust in the study findings. Using convenience sampling, we recruited participants who were top-cited scientists in the field of epidemiology, members of the Cochrane Bias Group and Cochrane Non-Randomized Studies Methods Group, authors of initiatives related to observational studies and corresponding authors of NRSIs. Through an online survey, we asked them to identify factors that they believe could influence the trustworthiness of NRSIs. We analyzed qualitative data using an inductive thematic approach. We first coded the responses, which were redefined into factors and grouped under themes. We summarized findings in frequencies and percentages.

**Results:** 130 participants out of 1488 contacted completed the survey. Of the 130 participants, 40 (31%) were methodologists and 61 (47%) had 21-40 years of experience in research. The level of expertise in NRSIs ranged from intermediate (35%) to advanced (30%) and expert (30%).We identified a total of 56 factors, with a median of 6 factors per participant (IQR 3; 9, range 0-20). We grouped the factors under 20 domains, when relevant, and eventually under eight overarching themes: Open Science (e.g., transparency, registration), Research Question (e.g., appropriate rationale and hypothesis), Study Methodology (e.g., study design, participants, statistical considerations), Data Source (e.g., quality), Findings and Interpretation (e.g., plausibility of effect estimate), Writing (e.g., appropriate writing), Oversight (e.g., investigators, journal), and Artificial Intelligence (e.g., no suspicion of use in writing or synthesis).

**Conclusions:** Our findings provide insight to gauge and improve the quality and uptake of NRSIs, with important implications for strengthening evidence-based decision-making in both research and practice.

## BACKGROUND

Generating reliable evidence to guide clinical decision-making is essential. Both randomized controlled trials (RCTs) and non-randomized (observational) studies of interventions play complementary roles in informing medical practice. While RCTs are considered the gold standard for assessing causal effects, non-randomized studies of interventions (NRSIs) are increasingly recognized as valuable, particularly with the growing availability of routinely collected data (1, 2) and the development of advanced design and analytical methods to mitigate biases, such as the target trial emulation framework (3).

Despite methodological advancements, concerns about the trustworthiness of non-randomized studies persist, largely due to challenges such as confounding and other biases (4). Significant efforts have been made to gauge systematically the presence and impact of biases, including risk of bias assessment tools, notably Risk of Bias in Non-Randomised Studies - of Interventions (ROBINS-I) tool (5). However, even when a study demonstrates minimal risk of bias on such tools, questions about its overall trustworthiness may remain.

Trustworthiness could be influenced by factors beyond the items traditionally included in risk of bias tools. These factors may include study-related characteristics, the research context, and external circumstances. Elements such as study transparency, protocol registration, data sharing, dataset registration, and replication of analyses may contribute significantly to how a study is perceived and trusted (6, 7). Identifying these factors is essential for gauging the credibility of non-randomized studies as they are interpreted and used in systematic reviews, and for improving their design to ensure that they provide reliable evidence for decision-making. In addition, these factors could be taken into account when evaluating the confidence in the results of systematic reviews that include NRSI.

This study aimed to identify factors, not covered in risk of bias assessment tools, that could influence the trustworthiness of NRSIs.

## METHODS

### Study design and setting

The study was a cross-sectional survey of experts. The protocol is registered on Open Science Framework (osf.io/9wdcz). The results from this study along with those from a scoping review will generate factors which will be sorted in a later step. We reported the study according to Consensus-Based Checklist for Reporting of Survey Studies (CROSS) (see Additional file 1: Table S1) (8).

We define trustworthiness as the proper, justified or rational trust in the study findings, i.e., if there is good reason to think that study findings are true or at least sufficiently close to the truth and that they are based on sufficient high-quality evidence (9). Although our definition includes internal validity, we consider trustworthiness more broadly, including factors not addressed in existing risk of bias assessment tools.

### Participants

We recruited participants who are methodologists and researchers with experience in observational studies, using convenience sampling. We invited researchers in the field of epidemiology, who are in the top 2% highly-cited scientists and who started publishing after 1984, identified from the author databases of standardized citation indicators (10).

Specifically, of 692 authors who are among the top-2% in citation impact for their career-long work with a primary or secondary field being epidemiology and who started publishing after 1984, we invited 677 authors with available contact information. In addition, we invited 491 members of the Cochrane Bias Group and Cochrane Non-Randomized Studies Methods Group, as well as 204 authors of initiatives related to the methodology (e.g., target trial emulation), risk of bias (e.g., ROBINS-I) and reporting (e.g., RECORD) of observational studies. The list of considered initiatives are listed in Additional file 1: Table S2. We also invited 116 corresponding authors of non-randomized studies of interventions published in six high impact-factor medical journals (Lancet, NEJM, JAMA, BMJ, Annals of Internal Medicine, JAMA Internal Medicine) in 2022-2023 (see Additional file 1: Table S3).

### Data collection

We invited selected scientists for participation via a standardized email to inform them of the study and invite them to participate in the online survey. Informed consent was obtained from study participants before the start of the survey. After one week of the initial contact (September 24, 2024), we sent two reminders; one-week apart. We pilot-tested the survey beforehand.

The invitations included a link to the survey (see Additional file 2), consisting of the informed consent and a set of questions on general characteristics such as gender, country of residence, field of expertise, years of experience, level of expertise in non-randomized studies of interventions, etc. Then, we asked participants to list factors, not covered in risk of bias assessment tools, that could influence their trust in the non-randomized study. These factors could pertain to the study itself, the field where it is performed, the circumstances under which it is performed, or any other factor that they may consider as influential. For each factor, the participants were asked to provide a brief definition and to specify whether it increases or decreases the trustworthiness of the study.

### Data analyses

We used thematic analysis (11), a process for encoding qualitative information. It involves free line-by-line coding of the responses, the organization of these free codes into related areas, and eventually the development of themes. These themes can be generated inductively from the raw information or deductively from theory or prior research (11, 12). In this study, one author manually coded the responses and a second author provided input, when needed. Then, we organized the open codes into factors based on similarities and differences between codes. Finally, we organized the factors into domains and overarching themes (broad categories covering main domains) using an inductive process. Throughout the coding process, the authors met regularly to discuss the codes, factors, domains and themes. Items that were explicitly mentioned in ROBINS-I (related to bias due to confounding, selection of participants into study, classification of interventions, deviations from intended interventions, missing data, measurement of outcomes, or selection of the reported result) (5), those that solely compared RCTs to NRSIs, and those related to the expertise of the reader were not considered. We report the factors in the ‘positive’ direction i.e., the presence of the factor may increase the trustworthiness of the study. We present examples of coded text excerpts in quoted italics. In addition, we conducted descriptive statistics using IBM SPSS Statistics V.21 to summarize our findings in a narrative and tabular formats. We reported the median (25^th^ percentile; 75^th^ percentile) or mean (standard deviation [SD]) for continuous outcomes and frequencies and percentages for categorical outcomes We performed post-hoc exploratory univariable logistic regression analyses to examine the association between participant characteristics and each of the identified themes. We report the crude odds ratios (ORs) with 95% confidence intervals (CIs). Confidentiality and anonymity of all participants was ensured by using only code numbers and de-identified data. The study was exempt from ethical review in accordance with French regulations.

## RESULTS

### Characteristics of the participants

Out of 1488 contacted, 132 participants responded to our survey, out of which 130 completed it, and 128 proposed at least one eligible factor.

Half of the participants were females (n=65, 50%), 70% were PhD holders (n=90) and 31% were methodologists (n=40). In addition, almost half of the participants had 21-40 years of experience in research (n=61, 47%), while 25% had 11-20 years of experience (n=32). The self-reported level of expertise in NRSIs ranged from intermediate (n=45, 35%) to advanced (n=39, 30%) and expert (n=39, 30%). Most participants had conducted NRSIs (n=78, 60%) or assessed them as part of evidence synthesis (n=103, 79%). We present a summary of the characteristics of the participants in Table 1.

**Table 1.** Characteristics of the participants (N=130).

| Characteristic |  | Frequency (Percentage) |
| --- | --- | --- |
| Age (N=122) |  | Mean 52.0 SD 14.1; Median 52.0 IQR (41.8; 63.0) |
| Gender (N=129) | Male | 63 (49%) |
|  | Female | 65 (50%) |
|  | Other | 1 (1%) |
| Country of residence (N=127)* | United States | 25 (20%) |
|  | United Kingdom | 21 (17%) |
|  | Canada | 18 (14%) |
|  | Australia | 13 (10%) |
|  | Germany | 9 (7%) |
|  | Denmark | 5 (4%) |
| Highest educational qualification | Master's Degree | 8 (6%) |
|  | Doctoral degree | 90 (69%) |
|  | Doctor of Medicine | 24 (19%) |
|  | Other | 8 (6%) |
| Current professional activity | Clinician | 13 (10%) |
|  | Methodologist | 40 (31%) |
|  | Epidemiologist | 29 (22%) |
|  | Editor | 3 (2%) |
|  | Systematic reviewer | 22 (17%) |
|  | Statistician | 14 (11%) |
|  | Other | 9 (7%) |
| Years of experience as a researcher | 1-5 years | 11 (9%) |
|  | 6-10 years | 14 (11%) |
|  | 11-20 years | 32 (25%) |
|  | 21-40 years | 61 (47%) |
|  | >40 years | 12 (9%) |
| **Level of expertise in NRSIs** | Novice | 1 (1%) |
|  | Beginner | 6 (5%) |
|  | Intermediate | 45 (35%) |
|  | Advanced | 39 (30%) |
|  | Expert | 39 (30%) |
| **Involvement in NRSIs** | Conducting NRSIs | 78 (60%) |
|  | Assessing NRSIs | 103 (79%) |
|  | Developing reporting guidelines | 35 (27%) |
|  | Other | 9 (7%) |

NRSI: non-randomized studies of interventions, SD: standard deviation, IQR: interquartile range
\*Only those with 5 counts or more are presented.

### Factors that could influence the trustworthiness of non-randomized studies of interventions

A total of 749 responses were proposed by the 130 participants. After discarding responses that were explicitly mentioned in ROBINS-I, those that solely compared RCTs to NRSIs and those that are related to the expertise of the reader (see Additional file 1: Table S 4), 674 reponses were recorded to code and analyze.

After coding the responses, we identified a total of 56 factors. We grouped the factors under 20 domains, when relevant, and eventually under eight overarching themes. The themes are: Open Science, Research Question, Study Methodology, Data Source, Findings and Interpretation, Writing, Oversight, and Artificial Intelligence. Figure 1 summarizes our findings. We describe each theme with examples in the following sections and in the codebook in Additional file 1: Table S5.

**Figure 1.**
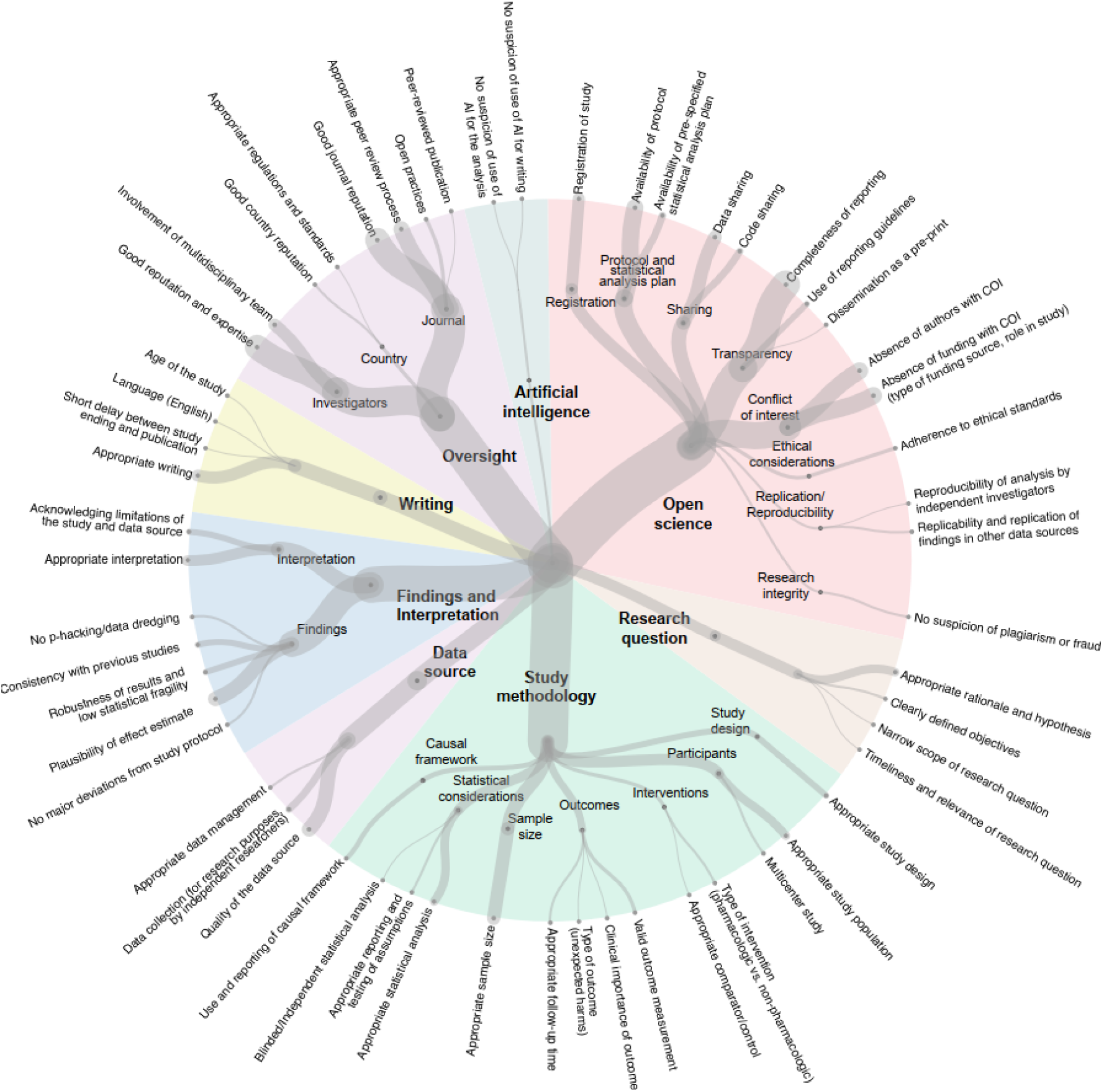
Factors that could influence the trustworthiness of non-randomized studies of interventions.

#### Open science

A total of seven domains were identified in the theme Open Science. One domain was registration of study where one respondent stated: “*prospective registration of the study protocol in an open registry increases my trust, as it indicates a more transparent process and less prone to bias*”. Another domain was protocol and statistical analysis plan, where one respondent stated: “*If no published protocol is available my trust in the reported results would decline*”. Sharing was also mentioned by many participants, one mentioning that “*A clear statement about data sharing, that looked as though it would be followed through on, would increase my trust in a study*” and another participant stating that “*if data is made available that would increase my trust*”. A frequently reported domain was transparency, which includes completeness of reporting (see Additional file 1: Table S6) and use of reporting guidelines. Moreover, domains such as conflict of interest and ethical considerations were also identified. Many respondents mentioned that the absence of funding with conflict of interest (which includes the type of funding source and role of the funding source in study) would increase their trust in the study. Another identified domain was replication and reproducibility. Respondents highlighted that “*studies that have been successfully replicated or validated by independent research groups tend to be viewed as more trustworthy”* and that *“the study’s ability to replicate or build upon existing findings enhance trust”.* The last domain identified was research integrity where participants underlined that suspicion of plagiarism or fraud would decrease their trust.

#### Research question

The theme research question encompasses several factors, with the most reported one being appropriate rationale and hypothesis. One participant stated “*Plausibility of the hypothesis (less plausible would decrease trust). This is based on evidence to support the mechanism of the hypothesis, and prior research that supports the hypothesis.”* Another respondent stated “*Studies of treatments that do not have a biologically sound basis for their effect are less trustworthy”*.

#### Study methodology

A total of seven domains were identified including study design, participants (e.g., representation of the true target population, multicenter study), interventions (e.g., appropriate comparator/control), outcomes (e.g., valid outcome measurement, clinical importance of outcomes and appropriate follow-up time), sample size, statistical considerations and causal framework (e.g., use and reporting of causal framework). Some participants mentioned that they would trust studies assessing a pharmacologic treatment, rather than a non-pharmacologic one. Participants also mentioned the type of outcome as a factor, for example if they were harms, they would trust it more; *“I would trust unknown harm outcome results unrelated to the disease under question more than known other outcomes because these ‘novel’ outcomes may be less susceptible to confounding by indication”*. Moreover, appropriate statistical analysis increases trust, in addition to appropriate reporting and testing of assumptions and blinded/independent statistical analysis. Lastly, the use and reporting of causal framework increases trust in studies. One respondent stated that *“study methodology includes target trial emulation - most appropriate methodology for observational study of interventions, [leads to] increased trust”*.

#### Data source

Respondents highlighted three factors related to data sources. One factor was the quality of the data source, stating: “*increased trust if the data source is high-quality and comprehensive”* and *“consistency in the dataset comes up irrespective of how the data is sliced and diced, raises the trustworthiness of the dataset/author group”.* Another factor was data collection. Some respondents mentioned that the use of data collected for research purposes increases their trust, stating *“registry databases or cohort studies, is often more suitable for causal inference than electronic medical record data or case-control studies”,* whereas another respondent believed otherwise *“a study planned from a large medical record data base might be more trustworthy than one from a clinical investigator with potential interest in a new intervention.”* Others highlighted the time of data collection *“if the data were collected prospectively, then this would likely increase my trust”.* The third factor was appropriate data management: *“Inappropriate data curation, transformation, linkage, and cleaning may decrease my trust in overall results”*.

#### Findings and interpretation

Two domains were identified. One domain was related to findings, where the main factor was the plausibility of the effect estimate, related to its size, and whether it is expected or ‘too good to be true’. The second domain was interpretation, where the main factor was appropriate interpretation, which includes absence of spin, having appropriate counter-arguments and comparison with other studies, and including the implications of the findings. One respondent stated *“Spin (decreases trust): where the discussion, conclusions and abstract of the result focus on positive messages, ignoring any nuance, uncertainty or concerns arising from the data.”*

#### Writing

This theme includes few factors. The main one is appropriate writing which was reported by many participants. It includes structure, consistency, clarity, stylistic quality, length, visualizations and citations. Another factor is the delay between study ending and publication, stating: “*short time increases my trust”.* In addition, two participants mentioned that they would trust studies that are more recently published more than older ones.

#### Oversight

Respondents mentioned factors related to oversight, i.e., overseeing the study. This includes investigators’ good reputation and expertise; the involvement of multidisciplinary team (including interest-holders) and collaborators. One respondent stated *“I tend to have more trust in studies published by multiple investigators from more than one institution.”* Another mentioned *“[increased trust when] attempts been made to meet DEI [diversity, equity, and inclusion]”.* The country where the study took place was another important domain, e.g, *“authors from countries associated with poor quality research (decrease trust)”.* Finally, journal reputation (i.e., metrics and properties of the journal) and the peer review process were mentioned: *“articles published in high-impact journal (Q1 and Q2 journals) will improve the confidence of the study”; “publication in a predatory journal will decrease my trust.”*; *“publication in a highly regarded journal with a rigorous peer review process may increase trust”* and *“open peer review (published reviewer names and comments) [increases my trust]”*.

#### Artificial intelligence

Four participants mentioned the use of artificial intelligence (AI) would affect their trust in the study. Participants mentioned the use of AI for the analysis or for writing, stating *“if the write up and/or analysis of the results used AI, then it would likely decrease my trust in the findings”*.

### Post-hoc exploratory analysis

A median of 6 factors were proposed per participant (IQR 3; 9, range 0-20), with a median of 4 themes (IQR 2;4, range 0-7). We present the number of experts proposing at least one factor per theme in Table 2 and an overview of the themes proposed per participant in Figure 2.

**Figure 2.**
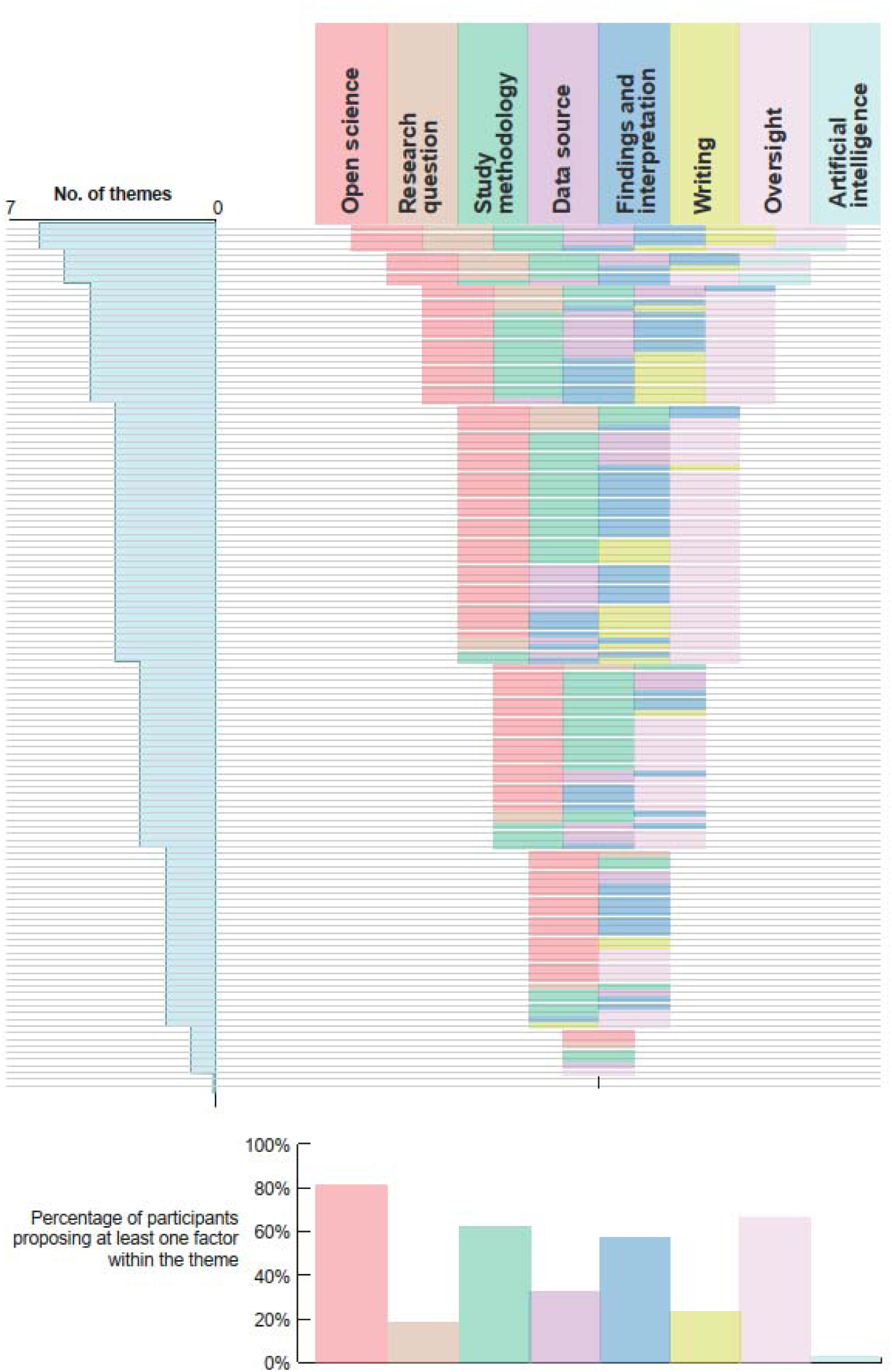
Overview of themes proposed per participant

**Table 2.** Reporting of themes.

| Reporting* | Open science | Research question | Study methodology | Data source | Findings and interpretation | Writing | Oversight | Artificial intelligence |
| --- | --- | --- | --- | --- | --- | --- | --- | --- |
| **One theme** | 2 | 1 | 2 | 1 | 0 | 0 | 1 | 0 |
| **Two themes** | 20 | 2 | 7 | 3 | 11 | 3 | 8 | 0 |
| **Three or more themes** | 84 | 21 | 72 | 38 | 64 | 27 | 78 | 4 |
| **Total** | **106** | **24** | **81** | **42** | **75** | **30** | **87** | **4** |
| **With other themes** |  |  |  |  |  |  |  |  |
| Open science | NA |  |  |  |  |  |  |  |
| Research question | 18 | NA |  |  |  |  |  |  |
| Study methodology | 66 | 18 | NA |  |  |  |  |  |
| Data source | 35 | 8 | 28 | NA |  |  |  |  |
| Findings and interpretation | 65 | 17 | 46 | 24 | NA |  |  |  |
| Writing | 27 | 7 | 19 | 6 | 20 | NA |  |  |
| Oversight | 75 | 15 | 58 | 31 | 54 | 26 | NA |  |
| Artificial intelligence | 4 | 3 | 4 | 2 | 4 | 1 | 3 | NA |

NA: not applicable
\*at least one factor reported by participant

Our exploratory analysis (Table 3) showed that each 10-year increase in age was associated with a 42% higher odds of proposing a factor under the theme Research Question (OR 1.42, 95% CI 1.01 to 2.00), but had 29% (OR 0.71, 95% CI 0.54 to 0.94) and 25% (OR 0.75, 95% CI 0.57 to 0.99) decreased odds for Study Methodology and Oversight, repectively. In addition, participants with more than 20 years of experience were less likely to propose factors under Study Methodology compared with those with 20 years of experience or less (OR 0.41, 95% CI 0.19 to 0.87). While those with advanced/expert level of expertise in NRSIs were less likely to propose factors under Open Science (OR 0.33, 95% CI 0.12 to 0.95) and Writing (OR 0.41, 95% CI 0.18 to 0.95). Similarly, participants who were involved in conducting NRSIs were less likely to propose factors under Open Science (OR 0.17, 95% CI 0.05 to 0.59), but more likely to propose ones under Findings and Interpretation (OR 2.52, 95% CI 1.23 to 5.19). Finally, participants who were involved in assessing NRSIs were less likely to propose factors under Research Question (OR 0.34, 95% CI 0.13 to 0.90). Additional details are reported in Additional file 1: Table S7.

**Table 3.** Association between participant characteristics and themes.

| Characteristic | Theme* |  |  |  |  |  |  |  |
| --- | --- | --- | --- | --- | --- | --- | --- | --- |
|  | Open science | Research question | Study methodology | Data source | Findings and interpretation | Writing | Oversight | Artificial intelligence |
| Age (per 10 years) | 0.99<br>(0.71 to 1.38) | 1.42<br>(1.01 to 2.00) | 0.71<br>(0.54 to 0.94) | 1.09<br>(0.83 to 1.42) | 1.02<br>(0.79 to 1.33) | 1.00<br>(0.75 to 1.35) | 0.75<br>(0.57 to 0.99) | 1.5<br>(0.71 to 3.16) |
| Years of experience (>20 years vs. ≤20 years) | 0.58<br>(0.23 to 1.48) | 1.38<br>(0.56 to 3.43) | 0.41<br>(0.19 to 0.87) | 1.23<br>(0.58 to 2.58) | 0.99<br>(0.49 to 1.99) | 1.03<br>(0.45 to 2.34) | 0.49<br>(0.23 to 1.06) | 2.4<br>(0.24 to 23.71) |
| Level of expertise (Advanced vs. Lower) | 0.33<br>(0.12 to 0.95) | 1.79<br>(0.69 to 4.68) | 0.7<br>(0.34 to 1.46) | 1.52<br>(0.71 to 3.28) | 1.14<br>(0.56 to 2.32) | 0.41<br>(0.18 to 0.95) | 0.53<br>(0.25 to 1.16) | 0.66<br>(0.09 to 4.82) |
| Involvement in conducting NRSIs (Yes vs. No) | 0.17<br>(0.05 to 0.59) | 1.42<br>(0.56 to 3.61) | 1.59<br>(0.77 to 3.27) | 1.52<br>(0.71 to 3.28) | 2.52<br>(1.23 to 5.19) | 0.59<br>(0.26 to 1.34) | 0.46<br>(0.21 to 1) | 2.04<br>(0.21 to 20.16) |
| Involvement in assessing NRSIs (Yes vs. No) | 1.77<br>(0.65 to 4.84) | 0.34<br>(0.13 to 0.90) | 0.97<br>(0.4 to 2.32) | 0.51<br>(0.22 to 1.23) | 0.62<br>(0.26 to 1.51) | 1.06<br>(0.39 to 2.94) | 1.86<br>(0.78 to 4.43) | 0.08<br>(0.01 to 0.79) |
| Involvement in development of reporting guidelines (Yes vs. No) | 2.07<br>(0.65 to 6.54) | 2.31<br>(0.92 to 5.85) | 0.46<br>(0.21 to 1.01) | 0.95<br>(0.41 to 2.18) | 1.14<br>(0.52 to 2.51) | 1.83<br>(0.77 to 4.39) | 0.78<br>(0.35 to 1.76) | 2.82<br>(0.38 to 20.82) |

\* Odds ratio (95% confidence interval); statistically significant results are highlighted

## DISCUSSION

In this study, we identified eight overarching themes, encompassing 20 domains and 56 factors that are not covered in the ROBINS-I risk of bias assessment tool, that may influence the trustworthiness of NRSIs. These themes correspond to different aspects of the research process. Some relate to specific stages of a study, such as the planning stage (e.g., registration, protocol and statistical analysis plan), conduct stage (e.g., study methodology) or reporting stage (e.g., interpretation, writing). Others relate to external factors such as oversight; investigators, journals, and country, which can evolve over time (e.g., peer review process, country regulations).

It is important to note that the identified factors also vary in terms of feasibility, complexity, and cost of implementation. For example, study registration and the availability of a protocol and statistical analysis plan are increasingly recognized as essential even for NRSI, not just for randomized trials (13). However, the implementation of these factors can be challenging, with ongoing debates around issues such as timing of protocol publication, availability of suitable platforms, and accessibility to the data (14). Data sharing is another complex issue. While the open science movement advocates for open data sharing, in the context of these studies, the primary data sources are often registries or electronic health records, which come with their own legal, ethical, and logistical restrictions (15, 16). Moreoever, factors like replication and replicability, might require their own set of expertise and cost to implement. In contrast, improving the completeness of reporting, adhering to reporting guidelines and appropriate writing are generally low-cost and, in theory - but not necessarily in practice - easier to implement (17–19). In addition, aritificial intelligence represents an emerging and probably disruptive technology, and perceptions that its use may decrease trust could change abruptly in the near future.

Several of the themes and factors we identified align with the ones existing in the literature such as Open Science, Study Methodology, Findings and Interpretation, Writing and Oversight. Open Science has been advocated broadly in the research community as a method to improving trustworthiness of research (20, 21). Open science practices support transparency, reproducibility, accessibility, traceability and verifiability of scientific research (9, 20, 22, 23), and have been shown to positively influence how credible a study is perceived to be (21, 24). In addition, consistent with literature on research waste, participants also highlighted the importance of methodological rigor, in addition to transparent reporting, which can compromise the trust in studies if inadequate (25, 26). Participants also emphasized the importance of how well results are contextualized, and whether the interpretation and conclusions align with findings. This highlights that trust of readers is not only influenced by how research is conducted, but also by how findings are interpreted and conclusions are communicated by the authors (27, 28). When conclusions are not well aligned with the findings, it may compromise the trustworthiness of the study as a whole. Lastly, we underscore the importance of the role of journals and investigators in trustworthiness of NRSIs. Studies have reported that peer review and other journal characteristics, such as impact factor, play a role in ensuring trustworthiness in science (29–31). Similarly, involving a multidisciplinary team, such as a statistician to assess the appropriateness of the proposed data analysis models, ensures research quality (20). Overall, the literature reinforces our finding that trustworthy research is not determined by a single factor, but rather by a combination of factors spanning various aspects of NRSIs and the wider environment where they are designed, conducted and published.

Our exploratory analysis suggests that perspectives on what makes NRSIs trustworthy vary by role and experience of the reader. Older participants were more likely to propose factors under Research Question but less likely to highlight Study Methodology and Oversight, perhaps reflecting a focus on broader scientific aims, rather than on the practical or technical details.

In contrast, participants conducting NRSIs proposed factors under Findings and Interpretation while downplaying Open Science and Writing, suggesting that the findings and their interpretation may take precedence over practices related to transparency and dissemination. Overall, the findings suggest that perceptions of trustworthiness may be shaped by the priorities of those with different research roles and levels of experience.

### Strengths and limitations

One strength of our study is the diverse characteristics of the participants. Their expertise spans various professional roles, countries, years of experience and levels of involvement in NRSIs. Another strength is the development of the survey questions in collaboration with several experts in the field. We also pilot-tested the survey, which allowed us to refine the questions for clarity and relevance. However, the key limitation of the study is the low response rate. This may be due to the fact that we requested from particiants substantial effort and cognitive involvement to complete the survey with meaningful information. Since we lack information about non-respondents, we cannot assess how they differ from those who participated. However, we believe that it is unlikely that we have missed a factor of importance not identified by the 130 participants, since the responses we obtained provide a substantial and diverse set of factors that should be further explored and validated in future research, specifically those proposed by participants who are involved in conducting or assessing NRSIs (e.g., Findings and Interpretation).

### Implications for practice and research

The findings of this study have several implications. While some factors may be indirectly related to risk of bias, our understanding of study trustworthiness - which has long been focused on the risk of bias - should be broadened to include additional factors not captured by existing assessment tools like ROBINS-I. Whether and how these factors are considered by stakeholders may be influenced by the complexity, resources, and various ethical and practical considerations. For researchers, the identified factors and themes can inform the design, conduct, and reporting of NRSIs, promoting greater transparency and methodological rigor.

For systematic reviewers, this work provides a foundation to appraise the included NRSIs and assess their trustworthiness when synthesizing evidence. Similarly, for peer reviewers and journal editors when reviewing reports of NRSIs. In addition, for decision-makers, the results underscore the importance of considering these factors when interpreting and using evidence from NRSIs to inform clinical and public health decisions. Finally, this study highlights that the quality of the data source itself is a critical factor, particularly in NRSIs. Although investigators often have limited control over the quality of the data they use, other stakeholders, such as data owners and institutions, can take active steps to enhance data quality. This underscores the shared responsibility across the research ecosystem for ensuring the trustworthiness of evidence.

Future research should further investigate the factors identified in this study and build on these findings to develop a prioritized list for assessing the trustworthiness of NRSIs, similar to the INSEPCT-SR tool currently in development for RCTs (32). In particular, it would be important to explore how participant characteristics - such as role, experience, and expertise - may influence perceptions of trustworthiness and the relative importance of different factors. This could inform the development of practical tools, such as checklists or frameworks to support researchers in conducting trustworthy NRSIs and to aid systematic reviewers, peer reviewers, journal editors and decision-makers in assessing the trustworthiness of NRSIs.

Finally, although our study focused on NRSIs, it is worth investigating how our findings might be transposable to other types of studies and fields, as many factors are not unique to either the study type or the field (e.g., oversight, artificial intelligence).

## CONCLUSIONS

In conclusion, we identified factors influencing the trustworthiness of non-randomized studies of interventions, organized into eight overarching themes ranging from open science (e.g., availability of protocol) and study methodology (e.g., appropriate statisitical anlaysis) to oversight (e.g., appropriate peer review process). The findings provide insight to improve the quality and uptake of NRSIs, with important implications for strengthening evidence-based decision-making in both research and practice.

### List of abbreviations

- CI: confidence interval
- CROSS: Consensus-Based Checklist for Reporting of Survey Studies
- NRSIs: non-randomized studies of interventions
- OR: odds ratio
- RCTs: randomized controlled trials
- ROBINS-I: Risk of Bias in Non-Randomised Studies - of Interventions

## DECLARATIONS

### Ethical approval and consent to participate

The study was exempt from ethical review in accordance with French regulations. Informed consent was obtained from all participants.

### Consent for publication

Not applicable

### Availability of data and materials

The datasets generated and/or analysed during the current study will be available in OSF (osf.io/9wdcz), upon study publication. Only coded datasets will be available as the original responses may include identifiable information.

## Authors’ contributions

SY, PR, IB conceptualized the study. SY contributed to the acquisition and analysis. SY and IB drafted the first version of the manuscript. JPAI, AH, RP, PR contributed to the design of the study, interpretation of data, and critically reviewed the manuscript. All authors read and approved the manuscript.

## Data Availability

Coded datasets will be available in OSF (osf.io/9wdcz), upon study publication. The original responses will not be available, as it includes identifiable information.

## Acknowledgements

We thank Dr. Astrid Chevance for her input on the survey. We thank the following experts for responding to the survey and agreeing to be acknowledged: Dr Adrian Barnett, Queensland University of Technology, Dr Afach Sivem, Epidemiology in Dermatology and Evaluation of Therapeutics (EpiDermE) - EA 7379 Université Paris Est Créteil (UPEC) Créteil France, Dr Ahmed Samir, Faculty of Physical Therapy, Cairo University, Giza, Egypt, Dr Aidan Cashin, Centre for Pain IMPACT, Neuroscience Research Australia, Randwick, Australia, Dr Amit Raval, Dr Amy Drahota, University of Portsmouth, Dr Andreas Lundh, Cochrane Denmark and Centre for Evidence-Based Medicine Odense, Dr An-Wen Chan, University of Toronto, Dr Areti Angeliki Veroniki, St. Michael’s Hospital, Unity Health Toronto and IHPME, University of Toronto, Dr Arianne Verhagen, Prof Musculoskeletal physiotherapy, Dr Ayat Ahmadi, Tehran University of Medical Sciences, Dr Barnaby Reeves, University of Bristol, Dr Belinda Burford, National Coalition of Independent Scholars, Dr Bianca Maria Maglia Orlandi, Post doctoral on Heart Institute of University of São Paulo, Dr Bilal Ahmad Khan, Office of Research, Innovation and Commercialization _ Khyber Medical University, Peshawar, Pakistan, Dr Bryan Chung, Dr Carole Lunny, University of British Columbia, Dr Caroline Sabin, UCL, Dr Celia Byrne, Uniformed Services University, Dr Christopher Schmid, Professor of Biostatistics, Brown University School of Public Health, Dr Claire Infante-Rivard, McGill University, Montréal, Canada, Dr Craig A. Umscheid, Agency for Healthcare Research and Quality, Dr Craig Ramsay, Aberdeen University, Dr David Colquhoun, University College London, Dr Dietrich Rothenbacher, Ulm University, Dr Emma Maund, University of Southampton, Dr Esha Ray Chaudhuri, Independent Health Equity researcher / Member, Healthcare Excellence Canada; WHO Global Learning; Dr Ebraheem Albazee, Kuwait Institute For Medical Specializations, Kuwait City, Kuwait, Dr Els Goetghebeur, Universiteit Gent, Dr Emma Sydenham, Cochrane Central Editorial Service, Dr Fan Mei, Chinese Evidence-based Medicine Center, MAGIC China Center, West China Hospital, Sichuan University, China, Dr Finbarr Allen, University College Cork, Ireland, Dr Gabriel Henrique de Oliveira Pokorny, Instituto de Patologia da Coluna, Dr Graziella Filippini, Cochrane Italy, Milan, Italy, Dr Harrison iHansford, School of Health Sciences, Faculty of Medicine and Health, UNSW Sydney, Australia, Dr Hugh Sharma Waddington, London School of Hygiene & Tropical Medicine, Dr Hywel Williams, Professor and Co-Director, Centre of Evidence-Based Dermatology, University of Nottingham, Dr Jennifer Evans, LSHTM, Dr Jesús López-Alcalde, Cochrane Associate Centre of Madrid / Cochrane Complementary Field - Switzerland Satellite, Dr Jim Schmeckenbecher, King’s College London, Dr Joanne Yaffe, University of Utah College of Social Work, Dr Joerg Meerpohl, Cochrane Germany, Dr Jorge Andrés Delgado-Ron, Faculty of Health Sciences, Simon Fraser University, Dr Jørn Wetterslev, Private Office, Tuborg Sundpark Denmark, Dr Jose A. Calvache, Universidad del Cauca, Colombia, Dr Kaley Hayes, Brown University, Dr Karen Robinson, Johns Hopkins University, Dr Katja Matthias, Faculty of Health Service, Catholic University of Applied Sciences of North Rhine-Westphalia, Germany, Dr Kieran Cooley, The Canadian College of Naturopathic Medicine, Dr Kim Madden, McMaster University, Dr Kristina Lindsley, IQVIA, Dr Lars Hemkens, University of Basel, Switzerland, Dr Lars Vatten, Norwegian University of Science and Technology, Dr Laurence Blanchard, London School of Hygiene & Tropical Medicine, Dr Liliya Eugenevna Ziganshina, Cochrane, Dr Lisa Hartling, University of Alberta, Dr Mariska M. G. Leeflang, Amsterdam UMC, University of Amsterdam, Dr Marta Roqué, Cochrane Iberoamerica - Sant Pau Research Institute (IR Sant Pau), Dr Martin Kulldorff, Dr Matthew Page, Monash University, Dr Maya Magdy Abdelwahab, Faculty of Medicine, Helwan University, Dr Melissa Sharp, RCSI University Of Medicine And Health Sciences, Dr Miranda Cumpston, Australian Living Evidence Collaboration, School of Public Health and Preventive Medicine, Dr Ole Olsen, The Research Unit for General Practice and Section of General Practice, Department of Public Health, University of Copenhagen, Denmark, Dr Onder Ergonul, Koç University School of Medicine, Dr Oyekola Oloyede, Sefako Makgatho Health Sciences University South Africa, Dr Peter Gøtzsche, Institute for Scientific Freedom, Dr Philip Greenland, MD, Northwestern University, Chicago, Dr Prof Tracy Merlin, School of Public Health, University of Adelaide, South Australia, Dr Ramkripa Raghavan, Nutrition Evidence Systematic Review Branch, Center for Nutrition Policy and Promotion, USDA, Dr Rebecca E Ryan, Cochrane Consumers and Communication group, Dr Regina Kunz, Div. of Clinical Epidemiology, University of Basel, Switzerland, Dr Robert M. Golub, MD, Northwestern University Feinberg School of Medicine, Chicago, Illinois, USA, Dr Roland Brian Büchter, Institute for Research in Operative Medicine (IFOM), Faculty of Health, School of Medicine, Witten/Herdecke University, Dr Rolf Groenwold, LUMC, Leiden, the Netherlands, Dr Sarah Tanveer, University of Maryland, Baltimore, Dr Sathish, Department of Orthopaedics, Government Medical College, India, Dr Silvia Minozzi, Laboratory of Methodology of Systematic Reviews and Guidelines production Mario Negri Pharmacological Research Institute IRCCS, Dr Tammy Hoffmann, Institute for Evidence-Based Healthcare, Bond University, Dr Tatyana Shamliyan, American College of Physicians, Dr Vanessa Jordan, University of Auckland, Dr Willi Sauerbrei, Faculty of Medicine and Medical Center - University of Freiburg, Dr Wong Chun Hoong, Hospital Kuala Lumpur, Dr Yen-Fu Chen, University of Warwick, Dr Yi Zhou, Beijing international center for statistical research, Peking Univerisity, Dr Yurkina Morales Femenias, Salud Pública., Dr Zachary Munn, HESRI (Health Evidence Synthesis, Recommendations and Impact), School of Public Health, University of Adelaide, Dr Zain Ali Nadeem, Allama Iqbal Medical College, Dr Farhad Shokraneh, University of Oxford, United Kingdom, and Dr Margaret Winker, World Association of Medical Editors

## ADDITIONAL FILES

**Additional file 1.** Tables S1-S11

- **Table S1.** Consensus-Based Checklist for Reporting of Survey Studies (CROSS)
- **Table S2.** List of initiatives related to non-randomized observational studies
- **Table S3.** Search strategy using Pubmed (Ovid)
- **Table S4.** Responses not considered
- **Table S5.** Codebook with detailed description of factors
- **Table S6.** Completeness of reporting (N=130)
- **Table S7.** Themes according to participant characteristics (N=130)

**Additional file 2.** Survey questions

