## Additional file 1 for "Factors influencing the trustworthiness of non-randomized studies of interventions: a survey of international experts"

### **Additional file 1.** Tables S1-S11

**Table S1.** Consensus-Based Checklist for Reporting of Survey Studies (CROSS)

**Table S2.** List of initiatives related to non-randomized observational studies

**Table S3.** Search strategy using Pubmed (Ovid)

**Table S4.** Responses not considered

**Table S5.** Codebook with detailed description of factors

**Table S6.** Completeness of reporting (N=130)

**Table S7.** Themes according to participant characteristics (N=130)

**Table S1.** Consensus-Based Checklist for Reporting of Survey Studies (CROSS)

| Section/topic | Item | Item description | Reported on page # |
|---|---|---|---|
| **Title and abstract** | | | |
| Title and abstract | 1a | State the word “survey” along with a commonly used term in title or abstract to introduce the study’s design. | p. 1 |
| | 1b | Provide an informative summary in the abstract, covering background, objectives, methods, findings/results, interpretation/discussion, and conclusions. | p. 2 |
| **Introduction** | | | |
| Background | 2 | Provide a background about the rationale of study, what has been previously done, and why this survey is needed. | p. 4 |
| Purpose/aim | 3 | Identify specific purposes, aims, goals, or objectives of the study. | p. 4 |
| **Methods** | | | |
| Study design | 4 | Specify the study design in the methods section with a commonly used term (e.g., cross-sectional or longitudinal). | p. 5 |
| Data collection methods | 5a | Describe the questionnaire (e.g., number of sections, number of questions, number and names of instruments used). | |
| | 5b | Describe all questionnaire instruments that were used in the survey to measure particular concepts. Report target population, reported validity and reliability information, scoring/classification procedure, and reference links (if any). | p. 5 |
| | 5c | Provide information on pretesting of the questionnaire, if performed (in the article or in an online supplement). Report the method of pretesting, number of times questionnaire was pre-tested, number and demographics of participants used for pretesting, and the level of similarity of demographics between pre-testing participants and sample population. | p. 6 |
| | 5d | Questionnaire if possible, should be fully provided (in the article, or as appendices or as an online supplement). | Appendix 3 |
| Sample characteristics | 6a | Describe the study population (i.e., background, locations, eligibility criteria for participant inclusion in survey, exclusion criteria). | p. 5 |
| | 6b | Describe the sampling techniques used (e.g., single stage or multistage sampling, simple random sampling, stratified sampling, cluster sampling, convenience sampling). Specify the locations of sample participants whenever clustered sampling was applied. | p. 5 |
| | 6c | Provide information on sample size, along with details of sample size calculation. | p. 5 |
| | 6d | Describe how representative the sample is of the study population (or target population if possible), particularly for population-based surveys. | N/A |

| | | | |
|---|---|---|---|
| Survey administration | 7a | Provide information on modes of questionnaire administration, including the type and number of contacts, the location where the survey was conducted (e.g., outpatient room or by use of online tools, such as SurveyMonkey). | p. 6 |
| | 7b | Provide information of survey's time frame, such as periods of recruitment, exposure, and follow-up days. | p. 6 |
| | 7c | Provide information on the entry process:<br>–>For non-web-based surveys, provide approaches to minimize human error in data entry.<br>–>For web-based surveys, provide approaches to prevent "multiple participation" of participants. | N/A |
| Study preparation | 8 | Describe any preparation process before conducting the survey (e.g., interviewers' training process, advertising the survey). | N/A |
| Ethical considerations | 9a | Provide information on ethical approval for the survey if obtained, including informed consent, institutional review board [IRB] approval, Helsinki declaration, and good clinical practice [GCP] declaration (as appropriate). | p. 7 |
| | 9b | Provide information about survey anonymity and confidentiality and describe what mechanisms were used to protect unauthorized access. | p. 7 |
| Statistical analysis | 10a | Describe statistical methods and analytical approach. Report the statistical software that was used for data analysis. | p. 6 |
| | 10b | Report any modification of variables used in the analysis, along with reference (if available). | N/A |
| | 10c | Report details about how missing data was handled. Include rate of missing items, missing data mechanism (i.e., missing completely at random [MCAR], missing at random [MAR] or missing not at random [MNAR]) and methods used to deal with missing data (e.g., multiple imputation). | p. 6 |
| | 10d | State how non-response error was addressed. | N/A |
| | 10e | For longitudinal surveys, state how loss to follow-up was addressed. | N/A |
| | 10f | Indicate whether any methods such as weighting of items or propensity scores have been used to adjust for non-representativeness of the sample. | N/A |
| | 10g | Describe any sensitivity analysis conducted. | N/A |
| **Results** | | | |
| Respondent characteristics | 11a | Report numbers of individuals at each stage of the study. Consider using a flow diagram, if possible. | p. 8 |
| | 11b | Provide reasons for non-participation at each stage, if possible. | N/A |
| | 11c | Report response rate, present the definition of response rate or the formula used to calculate response rate. | p. 8 |

| | | | |
|---|---|---|---|
| | 11d | Provide information to define how unique visitors are determined. Report number of unique visitors along with relevant proportions (e.g., view proportion, participation proportion, completion proportion). | p. 8 |
| Descriptive results | 12 | Provide characteristics of study participants, as well as information on potential confounders and assessed outcomes. | p. 8 |
| Main findings | 13a | Give unadjusted estimates and, if applicable, confounder-adjusted estimates along with 95% confidence intervals and p-values. | p. 14 |
| | 13b | For multivariable analysis, provide information on the model building process, model fit statistics, and model assumptions (as appropriate). | N/A |
| | 13c | Provide details about any sensitivity analysis performed. If there are considerable amount of missing data, report sensitivity analyses comparing the results of complete cases with that of the imputed dataset (if possible). | N/A |
| **Discussion** | | | |
| Limitations | 14 | Discuss the limitations of the study, considering sources of potential biases and imprecisions, such as non-representativeness of sample, study design, important uncontrolled confounders. | p. 17 |
| Interpretations | 15 | Give a cautious overall interpretation of results, based on potential biases and imprecisions and suggest areas for future research. | p. 16 |
| Generalizability | 16 | Discuss the external validity of the results. | p. 17 |
| **Other sections** | | | |
| Role of funding source | 17 | State whether any funding organization has had any roles in the survey’s design, implementation, and analysis. | p. 19 |
| Conflict of interest | 18 | Declare any potential conflict of interest. | p. 19 |
| Acknowledgements | 19 | Provide names of organizations/persons that are acknowledged along with their contribution to the research. | p. 20 |

**Table S2.** List of initiatives related to non-randomized observational studies

- STROBE: STrengthening the Reporting of OBservational studies in Epidemiology
- MOOSE: Reporting Guidelines for Meta-analyses of Observational Studies
- GRIPS: Guidelines for Reporting of Health Research involving Observational Data)
- STaRT-RWE: structured template for planning and reporting on the implementation of real world evidence studies
- TREND: Transparent Reporting of Evaluations with Nonrandomized Designs
- REPEAT: Reproducible Evidence: Practices to Enhance and Achieve Transparency
- GRACE: Good Research for Comparative Effectiveness
- RECORD: REporting of studies Conducted using Observational Routinely-collected Data
- GATHER: Guidelines for Accurate and Transparent Health Estimates Reporting
- TIDieR: template for intervention description and replication
- ROBINS-I: Risk Of Bias In Non-randomised Studies - of Interventions
- NOS: Newcastle-Ottawa Scale
- STRATOS initiative: STRengthening Analytical Thinking for Observational Studies
- OHDSI group: Observational Health Data Sciences and Informatics
- Target trial emulation:
  - Development of the TrAnsparent ReportinG of observational studies Emulating a Target trial (TARGET) guideline
  - Reporting of Observational Studies Explicitly Aiming to Emulate Randomized Trials: A Systematic Review

**Table S3.** Search strategy using Pubmed (Ovid)

We ran a search in PubMed to identify non-randomized studies of interventions published in 2022-2023 in: 1. Lancet, 2. NEJM, 3. JAMA, 4. BMJ, 5. Annals of Internal Medicine, and 6. JAMA Internal Medicine. The search strategy can be found in (Appendix 2).

| Search number | Search Details | Results |
|---|---|---|
| 1 | (("non random*"[Title/Abstract] OR "nonrandom*"[Title/Abstract]) AND ("study"[Title/Abstract] OR "studies"[Title/Abstract])) OR "NRSI"[Title/Abstract] OR (("non interven*"[Title/Abstract] OR "noninterven*"[Title/Abstract]) AND ("study"[Title/Abstract] OR "studies"[Title/Abstract])) OR (("cohort*"[Title/Abstract] OR "incidence"[Title/Abstract]) AND ("study"[Title/Abstract] OR "studies"[Title/Abstract] OR "analysis"[Title/Abstract])) OR "control groups"[MeSH Terms] OR "matched-pair analysis"[MeSH Terms] OR ("case*"[Title/Abstract] AND ("control*"[Title/Abstract] OR "comparison*"[Title/Abstract] OR "comparat*"[Title/Abstract] OR "compare*"[Title/Abstract] OR "referent*"[Title/Abstract] OR "compeer*"[Title/Abstract])) OR (("match*"[Title/Abstract] OR "control*"[Title/Abstract]) AND ("case*"[Title/Abstract] OR "group*"[Title/Abstract] OR "study"[Title/Abstract] OR "studies"[Title/Abstract] OR "pair*"[Title/Abstract] OR "control*"[Title/Abstract])) OR ("comparative study"[Publication Type] OR "epidemiologic studies"[MeSH Terms] OR "evaluation study"[Publication Type]) OR (("retrospective"[Title/Abstract] OR "prospective"[Title/Abstract] OR "longitudinal"[Title/Abstract] OR "follow up"[Title/Abstract] OR "followup"[Title/Abstract] OR "epidemiologic*"[Title/Abstract] OR "evaluation"[Title/Abstract]) AND ("study"[Title/Abstract] OR "studies"[Title/Abstract] OR "analysis"[Title/Abstract] OR "surveys"[Title/Abstract])) OR (("comparat*"[Title/Abstract] OR "comparison*"[Title/Abstract] OR "continuing"[Title/Abstract] OR "observational"[Title/Abstract]) AND ("study"[Title/Abstract] OR "studies"[Title/Abstract] OR "research"[Title/Abstract] OR "analysis"[Title/Abstract] OR "analyses"[Title/Abstract])) OR ("population based"[Title/Abstract] AND ("study"[Title/Abstract] OR "studies"[Title/Abstract] OR "analysis"[Title/Abstract] OR "analyses"[Title/Abstract])) OR ("historic*"[Title/Abstract] AND ("study"[Title/Abstract] OR "studies"[Title/Abstract] OR "analysis"[Title/Abstract] OR "analyses"[Title/Abstract])) OR "Time factors"[MeSH Terms] OR ("time"[Title/Abstract] AND "factors"[Title/Abstract]) OR ("cross section*"[Title/Abstract] OR "cross section*"[Title/Abstract] OR "Prevalence Studies"[Title/Abstract] OR "Prevalence study"[Title/Abstract] OR "prevalence analysis"[Title/Abstract] OR "prevalence analyses"[Title/Abstract] OR "survey"[Title/Abstract]) OR (("emulated"[Title/Abstract] OR "emulation"[Title/Abstract]) AND ("trial*"[Title/Abstract] OR "stud*"[Title/Abstract] OR "experiment*"[Title/Abstract])) OR (("target"[Title/Abstract] AND ("trial*"[Title/Abstract] OR "stud*"[Title/Abstract] OR "experiment*"[Title/Abstract])) OR (("reference"[Title/Abstract] OR "standard"[Title/Abstract]) AND ("trial*"[Title/Abstract] OR "stud*"[Title/Abstract] OR "experiment*"[Title/Abstract]))) OR ("Information Systems"[MeSH Terms] OR "Medical Records"[MeSH Terms] OR "Patient Discharge"[MeSH Terms] OR "Hospital Records"[MeSH Terms] OR "Health Services Research"[MeSH Terms] OR "insurance, health"[MeSH Terms]) OR "Registries"[MeSH Terms] OR ("database*"[Title/Abstract] OR "regist*"[Title/Abstract] OR "ehr"[Title/Abstract] OR "emr"[Title/Abstract] OR "secondary data"[Title/Abstract] OR "big data"[Title/Abstract]) OR (("health"[Title/Abstract] OR "health | 16,025,270 |

| | | |
|---|---|---|
| | care"[Title/Abstract] OR "healthcare"[Title/Abstract] OR "hospital*"[Title/Abstract] OR "patient*"[Title/Abstract] OR "physician*"[Title/Abstract] OR "medical"[Title/Abstract] OR "linked"[Title/Abstract] OR "surveillance"[Title/Abstract] OR "insurance"[Title/Abstract] OR "billing"[Title/Abstract] OR "admission"[Title/Abstract] OR "discharge"[Title/Abstract] OR "routine"[Title/Abstract] OR "routinely"[Title/Abstract] OR "claim*"[Title/Abstract] OR "administrative"[Title/Abstract] OR "registr*"[Title/Abstract] OR "real world"[Title/Abstract] OR "emergency department*"[Title/Abstract] OR "vigilance"[Title/Abstract]) AND ("record*"[Title/Abstract] OR "claim*"[Title/Abstract] OR "data"[Title/Abstract] OR "file*"[Title/Abstract] OR "registr*"[Title/Abstract] OR "evidence"[Title/Abstract] OR "information system*"[Title/Abstract] OR "summar*"[Title/Abstract] OR "card*"[Title/Abstract] OR "study"[Title/Abstract] OR "studies"[Title/Abstract])) | |
| 2 | drug therapy[MeSH Terms] OR "drug therapy"[Title/Abstract] OR "pharmacologic*"[Title/Abstract] OR "pharmacotherap*"[Title/Abstract] OR "pharmaco therap*"[Title/Abstract] OR "chemotherap*"[Title/Abstract] OR "chemo therap*"[Title/Abstract] OR "drug*"[Title/Abstract] OR "medication*"[Title/Abstract] OR "medicine*"[Title/Abstract] OR "dose*"[Title/Abstract] OR "dosing"[Title/Abstract] OR "intravenous*"[Title/Abstract] OR "oral"[Title/Abstract] OR "orally"[Title/Abstract] OR "subcutaneous*"[Title/Abstract] OR "subq"[Title/Abstract] OR "capsule*"[Title/Abstract] OR "tablet*"[Title/Abstract] OR "treatment*"[Title/Abstract] OR "pretreatment*"[Title/Abstract] OR "prescription*"[Title/Abstract] OR "agent*"[Title/Abstract] OR "regimen*"[Title/Abstract] OR "pharmaceutical preparations"[MeSH Terms] OR "controlled substances"[MeSH Terms] OR "dosage forms"[MeSH Terms] OR "drugs, essential"[MeSH Terms] OR "drugs, generic"[MeSH Terms] OR "drugs, investigational"[MeSH Terms] OR "nonprescription drugs"[MeSH Terms] OR "prescription drugs"[MeSH Terms] OR "prodrugs"[MeSH Terms] OR "substandard drugs"[MeSH Terms] OR "synthetic drugs"[MeSH Terms] OR "designer drugs"[MeSH Terms] OR "chemical actions and uses"[MeSH Terms] | 13,159,179 |
| 3 | #1 AND #2 | 7,104,491 |
| 4 | Review[Publication Type] OR "Systematic Review"[Publication Type] OR "Address"[Publication Type] OR "Biography"[Publication Type] OR "Bibliography"[Publication Type] OR "Case Reports"[Publication Type] OR "Clinical Conference"[Publication Type] OR "Comment"[Publication Type] OR "Congress"[Publication Type] OR "Editorial"[Publication Type] OR "Letter"[Publication Type] OR "Dictionary"[Publication Type] OR "Directory"[Publication Type] OR "Historical Article"[Publication Type] OR "Legal Case"[Publication Type] OR "Meta-Analysis"[Publication Type] OR "Guideline"[Publication Type] OR "News"[Publication Type] OR "Newspaper Article"[Publication Type] OR "Patient Education Handout"[Publication Type] OR "Legislation"[Publication Type] OR "Lecture"[Publication Type] OR "Video-Audio Media"[Publication Type] OR "Webcast"[Publication Type] OR "Portrait"[Publication Type] OR "clinical trial, phase i"[Publication Type] OR "clinical trial, phase ii"[Publication Type] OR "Randomized Controlled Trial"[Publication Type] | 9,081,204 |
| 5 | Animals[MeSH Terms] NOT "Humans"[MeSH Terms] | 5,204,345 |
| 6 | #4 OR #5 | 13,959,964 |
| 7 | #3 NOT #6 | 4,331,807 |
| 8 | #7 AND "English"[Language] | 3,988,531 |
| 9 | lancet london england[Journal] OR "The New England journal of medicine"[Journal] OR "JAMA"[Journal] OR "British medical journal"[Journal] OR "Annals of Internal Medicine"[Journal] OR "jama internal medicine"[Journal] | 449,632 |
| 10 | #9 AND #8 | 547 |

**Table S4.** Responses not considered

Responses that were not considered included those that solely compared RCTs to NRSIs, those that are related to the expertise of the reader (i.e., prior beliefs/gut feeling, unfamiliarity with the field of study) and those that are covered in ROBINS-I. We report the list of proposed factors for the latter according to the risk of bias domains of ROBINS-I in the table below.

| **Risk of bias domain** | **Proposed factor** | **n** |
|---|---|---|
| Bias due to confounding | Appropriate adjustment for confounding | 18 |
| | Group comparability of baseline characteristics | 4 |
| | Residual confounding | 3 |
| | Confounding bias | 1 |
| Bias in selection of participants into the study | Selection bias | 1 |
| | Immortal bias | 1 |
| | Selection bias because of lack of randomization | 1 |
| Bias in selection of the reported result | Publication bias | 2 |
| | Non-selective reporting of results | 1 |
| Bias in classification of interventions | Misclassification of exposures | 2 |
| Bias due to deviations from intended intervention | Contamination or co-interventions | 1 |
| Bias due missing data | Bias from missing data | 1 |
| | Handling of missing data | 4 |
| | Amount of loss to follow-up | 4 |
| Bias in measurement of outcomes | Misclassification of outcomes | 2 |
| | Measurement bias | 1 |
| Bias in reporting | Publication bias | 2 |
| | Reporting bias | 1 |
| Risk of bias | Internal validity | 2 |
| | Likelihood of biases | 1 |

**Table S5.** Codebook with detailed description of factors

| **Theme/** Subtheme | Factor | n | Definitions/*Text excerpt examples* |
|---|---|---|---|
| **Open science** | | | |
| | Open science | 2 | Refers to the general concept of open sicence.<br><br>*"My trust would increase if the study was following practices of open research"* |
| Registration | Registration of study | 25 | Refers to registration of the study protocol. Respondents also highlighted that it should be done prosepctively.<br><br>*"Availability of a protocol that appears to have been developed/published or at least key design elements made available on a trial registry prior to the study starting. Knowing that the design decisions, objectives, outcomes etc were prespecified prior to the study starting would increase my trust."*<br><br>*"Registration of a detailed protocol in a publicly accessible database before the study was started would increase my trust in the study findings."*<br><br>*"Registered or there is a publicly accessible / published protocol [means] more trustworthy"*<br><br>*"Increase trust if registered prospectively"* |
| Protocol and statistical analysis plan | Availability of protocol | 37 | Refers to having a an accessible protocol. Respondents detailed that the protocol should be pre-planned (before any analysis) and publicly available.<br><br>*"Existence of a protocol published prior to the analysis being conducted, leads to greatly increased trust in the results"*<br><br>*"Having a detailed registered protocol with the methods of choosing the primary and secondary outcomes"*<br><br>*"[presence of] published protocol would indicate increased trustworthiness."* |

| | | | |
|---|---|---|---|
| | Availability of pre-specified statistical analysis plan | 15 | Refers to having an accessible planned statistical analysis plan (SAP). Respondents also specified that the SAP should be registered.<br>*"A detailed prospectively registered protocol and pre-specified SAP (to define the primary and secondary outcomes, their derivation from measured variables, methods for managing protocol deviations and missing data, planned subgroup and sensitivity analyses, and their interpretation). Its existence increases trust"*<br>*"Availability of a SAP [statistical analysis plan] that appears to have been prespecified would increase my trust, due to the increased confidence that the data was not cherry-picked after the trial/analysis ended."*<br>*"Did the authors create a causal diagram. Ideally this would be in the protocol. A causal diagram increases my confidence in the results. It shows that the authors have spent some time thinking about the variables and pathways, rather than just rushing into the analysis."* |
| Sharing | Data sharing | 29 | Refers to having accessible datasets i.e., open source data. Respondents also highlighted having a data sharing statement and authors being responsive to requests for sharing their data.<br>*"data availability/sharing - increased trust if dataset available"*<br>*"A clear statement about data sharing, that looks as though it would be followed through on, would increase my trust in a study."* |
| | Code sharing | 16 | Refers to having accessible codes i.e., open source code.<br>*"Studies that make their data and analysis code openly available for scrutiny and replication tend to inspire more confidence in their findings."*<br>*"Provision of a detailed appendix providing definitions for all study variables. Seeing that the authors clearly define all study variables in depth allows replicability. Ideally they also provide statistical code. [increase trust]"* |

| | | | |
|---|---|---|---|
| | | | *"Are the data and code publicly available. Researchers who are willing to share their data and code are generally likely to be thorough and careful. This is important for transparency, but it also gives me increased confidence that the results are not fabricated or hacked."* |
| Transparency | Completeness of reporting | 59 | Includes adequate reporting of study design, study methodology, sample size calculation, statistical analysis, participant recruitment (e.g., refusals, flowchart), results, study period, description of interventions, treatment assignment, outcomes and measurement tools, data collection, data sources and their quality, missing data, and study variables. It also includes overall transparency in reporting, presence of declaration statement, disclosure of the funding source, reporting deviations from protocol, reporting of contact details of investigators, and availability of appendices.<br><br>*"Clear and understandable presentation of the study design. This would increase trust, as I could understand what was actually done."*<br><br>*"Detailed reporting of methodological and statistical issues: Detailed reporting of analyses, potential confounders, etc in a paper would increase my trust"*<br><br>*"Sample size for the study calculated, reported and met for the analysis to be conducted, increases my trust"*<br><br>*"Completeness in reporting. I react poorly to incomplete or sloppy reporting. When I am looking for important specific and detailed enough information I expect to find it and if not, this raises my doubts (e.g., (1) how many controls (for instance in a CC study) were first selected and how many in this selection participated; (2) e.g., when and how were the questionnaires administered for the different study groups); e.g., in a longitudinal study: how many were originally selected as eligible and how many participated; at each follow-up, how many were lost and how many were analyzed."*<br><br>*"Little reporting of the results data would decrease my level of trust in the study. for instance, failure to report the data (means and variances, other relevant basic descriptive statistics) and only reporting the results of statistical tests would lead me to very low levels of trust in the study"* |

| | | | |
|---|---|---|---|
| | | | *"The time period that individuals were observed should be reported. Studies that do not report when the study took place seem less trustworthy."*<br><br>*"My trust would decrease if there was ambiguity around what the intervention was (quality of reporting poor, i.e. not following the TIDieR checklist), because this often feels like it's associated with other issues in the study."*<br><br>*"Poor detail about recruitment or sampling, how the intervention was delivered or the outcomes were assessed; decreases my trust"*<br><br>*"Complete reporting of data collection procedures increases my level of trust"*<br><br>*"Increase trust: conflicts of interest are fully disclosed in the published documents."*<br><br>*"Full disclosure of funding sources and any conflicts of interest improves transparency and trust."*<br><br>*"Reporting quality: I always trust papers more when authors follow nicely all reporting guidelines, and have a rather detailed reporting on the methodology, including deviations from protocol …"*<br><br>*"Decrease: If a study has incomplete address/contact details of investigator(s)"*<br><br>*"Availability of supplementary materials for any additional data that could not be presented in the manuscript increases reliability"* |
| | Use of reporting guidelines | 13 | Refers to the use of standardized reporting guidelines.<br><br>*"following relevant reporting guidelines increases trust"* |
| | Dissemination as pre-print | 1 | Refers to publishing the article/results as pre-print.<br><br>*"Did the authors post the results as a preprint. This would increase my trust. It would show that the authors are interested in getting feedback about their research and happy to openly share their results, indicating their own confidence in their results."* |

| | | | |
|---|---|---|---|
| Conflict of interest | Absence of author conflict of interest | 36 | Refers to absence of declared conflict of interest (COI) and of hidden undeclared COIs.<br><br>*"Conflicts of interest - stronger/more substantive conflicts of interest to the study finding decreases trust."*<br><br>*"Conflicts of interest among the author group, for example, surgical intervention done by patent holder would decrease my trust in results."*<br><br>*"Trust is decreased by failure to disclose potential or hidden conflict of interest"*<br><br>*"Potential financial conflict of interest not included in the disclosures (This would decrease trust)."* |
| | Absence of funding conflict of interest (type of funding source, role in study) | 37 | Refers to the absence of funding conflict of interest which includes the type of the funding source, in addition to the role of the funding source in the study.<br><br>*"Conflicted funding (decrease trust): Studies conducted or sponsored by organisations with a financial conflict of interest in the topic are very low in trust."*<br><br>*"Industry-funded studies might be viewed with more skepticism, while studies funded by independent sources may be seen as more trustworthy - generally decreases trust if there are potential conflicts or industry funding; increases if funding is from independent sources with no apparent conflicts."*<br><br>*"My trust would decrease if the work was funded or conducted by the manufacturers of the intervention"*<br><br>*"Level of involvement of the sponsor in the design, execution, and analysis of the study, and choice of journal for submission. (Higher level would decrease trust). Any involvement of the sponsor in any of these activities would decrease trust."* |
| Ethical considerations | Adherence to ethical standards (ethical approval, informed consent) | 8 | Refers to obtaining ethical approval (or waiver) and informed consent from participants when relevant. |

| | | | |
|---|---|---|---|
| | | | *"If the study lacks evidence of appropriate ethical oversight or if informed consent was not properly obtained from participants, this raises concerns about trustworthiness."*<br><br>*"ethics approval that predates study start date increases trust"* |
| Replication/Reproducibility | Reproducibility of analysis by independent investigators | 2 | Refers to analysis being reproduced or replicated by independent investigators (e.g., independent research groups)<br><br>*"Studies that have been successfully replicated or validated by independent research groups tend to be viewed as more trustworthy…"*<br><br>*"[Trust] decreased: Only published from ones with commercial interest and no independent research organization replicating the efforts"* |
| | Replicability and replication of findings in other data sources (across different populations, settings) | 5 | Refers to replicability and replication of findings in other data sources including different populations and different setting.<br><br>*"the study must be repeated in 2 or more independent samples; would increase trust"*<br><br>*"[Trust] increased: Replications of findings in different databases with consistency"* |
| Research integrity | Peer-reviewed publication | 8 | Refers to studies that have been peer-reviewed.<br><br>*"Existence of a peer-reviewed publication. Reports that are not peer-reviewed have not passed through an important amount of screening by experts. Therefore, I am less willing to trust the results of a report if it has not been published in a peer-reviewed venue."* |
| | No suspicion of plagiarism or fraud | 4 | Refers to suspicion/evidence of plagiarism or fraud in the study.<br><br>*"Evidence of potential fraudulent behavior, e.g., unusual numerical values such as similar patient characteristics as other studies; would decrease trustworthiness."* |

| | | | |
|---|---|---|---|
| | | | *"No evidence of figure manipulation... Tables and Figures are not reproduced from another source (eg, a screenshot, a different font, etc) unless labeled as such (plagiarism)"* |
| **Research question** | | | |
| | Appropriate rationale and hypothesis | 17 | Refers to an appropriate rationale and hypothesis, including mechanistic plausibility and being supported by the literature.<br><br>*"How well the rationale for the study is developed and based on other knowledge, increases trust"*<br><br>*"Plausibility of the hypothesis. This is based on evidence to support the mechanism of the hypothesis, and prior research that supports the hypothesis. (Less plausible would decrease trust.)"* |
| | Clearly defined objectives | 4 | Refers to explicitly reporting the objectives of the study.<br><br>*"Clear statement of purpose of the study [increases trust]"* |
| | Narrow scope of research question | 3 | Refers to the specificity of the research question.<br><br>*"A more focused study leads to (slightly) greater trust, since fishing for an interesting result is less of a concern"* |
| | Timeliness and relevance of research question | 1 | Refers to providing timely evidence on an important question.<br><br>*"Studies addressing timely and highly relevant questions may be viewed more favorably"* |
| **Study methodology** | | | |
| Study design | Appropriate study design | 13 | Refers to the choice of design and justification.<br><br>*"The type of study and design is essential: prospective, cohort [increases trust]"* |

| | | | |
|---|---|---|---|
| | | | *"Lack of rationale for an NRSI: it may decrease [trust]"*<br><br>*"The purpose of the research is an important aspect to consider. There are several reasons for conducting a NRSI, so the choice of this design should be carefully justified."* |
| Participants | Appropriate study population | 19 | Refers to having a representative sample.<br><br>*"Evidence that it [sample] is representative of the underlying population, increases confidence"*<br><br>*"More representative sample give greater trust."* |
| | Multicenter study | 6 | Refers to having a multicenter study.<br><br>*"Increase my trust: multicentric observational studies demonstrate the intervention can be done by more than one institution"*<br><br>*"Single center study few participants included decreases trustfulness"* |
| Interventions | Type of intervention (pharmacologic vs. non-pharmacologic) | 4 | Refers to having the type of the intervention. Respondents noted to trust studies assessing pharmacologic interventions more than those assessing other types of interventions such as complementary or alternative therapies.<br><br>*"Evaluation of non-pharmacological or complementary/alternative therapies decreases my trust compared to pharmacological treatments, as I am more prone to think that there are fewer guarantees that the study processes are rigorous"* |
| | Appropriate comparator/control | 3 | Refers to the use of appropriate comparator and controls.<br><br>*"If the study did not include appropriate controls, I don't think the study has any scientific value"* |

| | | | |
|---|---|---|---|
| | | | *"Use of Historical Controls: Comparing current intervention participants with controls from the past (historical controls) can be problematic if other changes over time (like improvements in care) explain the differences in outcomes."* |
| Outcomes | Valid outcome measurement | 8 | Refers to the use of consistent and valid outcome measurement.<br><br>*"Outcome selection must be aligned with clinical dilemmas and study questions. Outcome ascertainment must be validated, e.g. mortality cause from verbal autopsy without proper histopathology."*<br><br>*"My trust may decrease if the outcome measurement scale is reported differently to other reports, e.g. the minimum/maximum scores don't align with what you would expect for that scale."*<br><br>*"Does the outcome measurement actually measure the outcome of interest as opposed to using a proxy outcome for an actual outcome of interest?"* |
| | Clinical importance of outcome | 3 | Refers to the clinical importance of outcomes.<br><br>*"Decrease trust: Focus on clinically less important outcomes (or other less relevant aspects of the study);"*<br><br>*"Increase trust: outcomes that are highly valued by people"* |
| | Type of outcome (i.e., unexpected harms) | 3 | Refers to the type of the outcome. Respondents noted safety, long-term, objective outcomes would increase trsut.<br><br>*"I would trust unknown harm outcome results unrelated to the disease under question more than known other outcomes because these 'novel' outcomes may be less susceptible to confounding by indication".* |
| | Appropriate follow-up time | 5 | Refers to having appropriate follow-up time for outcome assessment.<br><br>*"Increase trust: long-term follow-up results for chronic conditions."* |

| | | | |
|---|---|---|---|
| | | | *"Have the authors chosen a follow-up period that is adequate to show the effect"* |
| Sample size | Appropriate sample size | 29 | Refers to studies with a large sample size.<br><br>*"Number of included patients: large number indicates more trust."*<br><br>*"Sample size: I would be more likely to trust a study with a very large sample (ie 500 or more)"* |
| Statistical considerations | Appropriate statistical analysis | 18 | Refers to applying appropriate statistical analysis.<br><br>*"Statistical analysis: Inappropriate statistical analysis is a fatal flaw, removing all trust in the study."*<br><br>*"Inaccurate analysis method will impact the outcome of the investigation, leading to poor quality paper and reduce the confidence of the study."* |
| | Appropriate reporting and testing of assumptions | 4 | Refers to appropriate reporting and testing of assumptions.<br><br>*"If the assumptions of the used statistical tests are not tested my trust in the observed 'effect' will decline."*<br><br>*"Explicit assumptions when assessing causality: without being clear about the assumptions made, many observational research claims causality far beyond their limitations (including causal language and action recommendations)."* |
| | Blinded/Independent statistical analysis | 3 | Refers to conducting statistical analysis that is blinded and independent. 3<br><br>*"Lower trust if the analysis was not done in a blinded fashion."*<br><br>*"Blinded statistical analysis: would increase my trust because it reduces the degrees of freedom in the analysis."*<br><br>*"independent analysis [increases]"* |

| | | | |
|---|---|---|---|
| Causal framework | Use and reporting of causal framework (including diagrams and estimands) | 11 | Refers to the use and reporting of causal framework, including directed acyclic graphs, and estimands.<br><br>*"Explicit causal intentions: leads to higher trust. If the authors are aiming to estimate a causal estimate then they tend to be more explicit about the estimand etc."*<br><br>*"Good methodology and study design that is thoughtful and attempts to use methods to emulate an RCT are primarily the methods I would use to judge other studies"* |
| **Data source** | | | |
| | Quality of the data source (fit-for-purpose and completeness) | 30 | Refers to the quality of the data source, including the fitness-for-purpose and completeness.<br><br>*"Quality of the dataset, if deemed to be of higher quality, then this would increase my trust in the study"*<br><br>*"Quality of dataset: population-based, complete data etc would increase trust"*<br><br>*"The source, scale and sample size of data and their fit with the research question."*<br><br>*"Study dataset (is the data set known to be a valid source of observational data) - increase trust"*<br><br>*"Increases trust if the data source is high-quality and comprehensive"* |
| | Data collection (e.g., for research purposes/independent researcher vs. routinely collected data) | 16 | Refers to the use of data collected by independent researchers and for research purposes.<br><br>*"increased if reasonable justification for data collected and appropriate methods for collecting quality data (and minimizing missing data)"*<br><br>*"greater trust in the data collection process to studies that mention that data was collected in the field by independent researchers and that data was entered in their system and verified by another person"* |

| | | | |
|---|---|---|---|
| | | | *"Increase trust: data from national registries or electronic health records"*<br><br>*"Strong confidence in prospectively collected datasets, e.g. registry databases or cohort studies, is often more suitable for causal inference than electronic medical record data or case-control studies."*<br><br>*"Studies based on individual data vs aggregate data/ecological studies, the former is more trustworthy"*<br><br>*"Purpose built research dataset or healthcare administrative database, and reason/ accuracy of data collection at source. More trust if dedicated database with pre-specified variables rather than routinely collected data for other purposes (insurance etc.)"*<br><br>*"Decrease trust: reliance on routinely collected data within a clinical practice or institutional setting, such as a health care network."* |
| | Appropriate data management (validation methods, cleaning) | 6 | Refers to appropriate data management, including validation methods and cleaning.<br><br>"*Inappropriate data curation, transformation, linkage, and cleaning may decrease my trust in overall results."*<br><br>*"Thorough reporting of data collection and validation methofds, cleaning and handling."*<br><br>*"The conduct and reporting of the initial data analysis for checking of the data quality (and errors) and understanding the scope of the actual study."* |
| **Findings and Interpretation** | | | |
| Findings | No major deviations from study protocol | 6 | Refers to not having major deviations from study protocol.<br><br>*"Studies that have pre-registered their protocols and adhered to them demonstrate a commitment to transparency"* |

| | | | |
|---|---|---|---|
| | | | *"Did they have a protocol and did they stick to the protocol. Did they report any departures from the protocol. The protocol is the number 1 factor for me for increasing confidence as so many studies are p-hacked. I want to know that they had a plan and stuck closely to it."* |
| | Plausibility of effect estimate (size, unexpected, too good to be true) | 30 | Refers to the plausibility of effect estimate relating to its size or whether its unexpected or too good to be true.<br><br>*"Effect size and plausibility - decreases trust if very large or implausible effect sizes"*<br><br>*"If data and results are too good to be true"* |
| | Robustness of results and low statistical fragility | 14 | Refers to the robustness of results and low statistical fragility.<br><br>*"Conducting clear and well-reasoned sensitivity analyses - increase"*<br><br>*"Study size, statistical fragility: larger studies and higher event rates are more trustworthy than smaller studies (all else equal)"*<br><br>*"Robustness and sensitivity analyses increase confidence"* |
| | Consistency with previous studies | 12 | Refers to having consistency of results with previous studies.<br><br>*"Corroboration of already published results… Consistency of results increases my trust in results of NRSIs."*<br><br>*"Findings that align with existing evidence or biological mechanisms may be viewed as more credible."* |
| | No p-hacking/data dredging | 6 | Refers to not p-hacking nor data dredging.<br><br>*"Lack of Pre-Specified Analysis Plan - decrease trust: If the study does not have a pre-specified analysis plan (i.e., how data will be analyzed and which outcomes will be assessed), it raises concerns and increases the likelihood of data dredging or p-* |

| | | | |
|---|---|---|---|
| | | | *hacking, where researchers may selectively explore different analyses until they find significant results."*<br><br>*"Were there a limited number of hypotheses that were tested, or was it a data dredging exercise. This is another p-hacking concern... Papers will lots of results tend to decrease my trust."* |
| Interpretation | Appropriate interpretation (no spin, appropriate counter-arguments, comparison with other studies, implications of findings) | 24 | Refers to appropriate interpretation of findings, including absence of spin, appropriate counter-arguments, comparison with other studies, and implications of findings.<br><br>*"If results are overstated and generalizations are made in the discussion which are not supported by the analysis, then this decreases my trust."*<br><br>*"Inadequate or improper citations/referencing decreases the trust in the non-randomized study of interventions. Examples of this include: no referencing systematic reviews or evidence synthesis in the field in neither the introduction nor discussion, no mention of counter-arguments whatsoever, inadequate framing of findings with the larger research landscape in the discussion, too brief of a limitations section or a limitations section with a large amount of spin, and overall a very brief reference list."* |
| | Acknowledging limitations of the study and data source | 14 | Refers to acknowledging the limitations of the study and the data source.<br><br>*"Less acknowledgement of limitations provided, the less trustworthy."*<br><br>*"The source of the data and the description of the authors of the limitations of that source. If the authors do not acknowledge the limitations and challenges of using for example electronic health records in a specific context, this decreases my trust."* |
| **Writing** | | | |
| | Appropriate writing (structure, consistency, clarity, stylistic quality, length, visualizations and citations) | 26 | Refers to appropriate writing including structure, consistency, clarity, stylistic quality, length, visualizations and citations.<br><br>*"Messy papers: decrease my trust"* |

| | | | |
|---|---|---|---|
| | | | *"Consistency of results:… inconsistency can take several different forms/types, but typical sources of inconsistency are: Results are inconsistent between tables/figures and the results section, e.g. a positive effect is a negative effect in the table. Discussion and results sections are inconsistent… For example, the results section reports no effect of the intervention, while the discussion praises the effectiveness of the intervention. Here, the discussion was - apparently - written before the results were available and not really adjusted afterwards."*<br><br>*"Clarity of writing. Clear and organized writing throughout the report will increase my trust."*<br><br>*"The quality of the writing is a big influencer for me. If the article is well written, the methods are clear to follow, and the results are presented in a straightforward manner then I have more trust in the findings. Part of this is that the authors correctly use terminology that is used in the field, both clinically and methodologically."*<br><br>*"The length. I find that very long papers are often not as easy to understand and very short ones very incomplete... Anything missing or presented in a cryptic way reduces credibility."*<br><br>*"Visualization of critical parts of the data in the paper and/or the supplemental material: more trustworthy"*<br><br>*"Lack of clear numerical results, use of complicated 'beautiful' graphs - decreases my trust"*<br><br>*"The set of references listed: whether the solid literature has been referenced and whether the references are sufficiently up to date and appear themselves trustworthy."* |
| | Short delay between study ending and publication | 2 | Refers to having a short delay between study ending and publication.<br><br>*"If this time is relatively short, this would increase my trust. If the time was very long, it would increase my suspicion that perhaps the study had issues that meant it needed to be submitted to various journals before finally being accepted, or that the results* |

| | | | |
|---|---|---|---|
| | | | *needed to be reworked/reanalyzed to come up with something that could be published."* |
| | Language (English) | 1 | Refers to study being published in English language.<br><br>*"Publication in a non-English language decreases my trust"* |
| | Age of the study | 2 | Refers to the recency of the study.<br><br>*"The time of publication, the more recent the more is trusted"* |
| **Oversight** | | | |
| Investigators | Good reputation and expertise | 52 | Refers to the reputation and expertise of the investigators, including having a track-record of publications.<br><br>*"Who is leading the research and the reputation of the research centre they are in. If the research centre is well-recognized in the field, it helps to increase my trust in the quality of the research."*<br><br>*"A prestigious institution or author might increase trust in the result."*<br><br>*"Author experience, affiliation: if track-record and reputable institution: increase trust"*<br><br>*"History of papers retracted because of pervasive errors, or requiring later publication of major corrections. (This would decrease trust). Even if this does not represent deliberate falsification, it points to a sloppiness in research skills."*<br><br>*"Reputation and track record of the research team: The credibility of the researchers conducting the study can influence trust. A history of high-quality research, academic integrity, and expertise in the specific field may increase trust, while past controversies or retractions could decrease it."*<br><br>*"Investigators: number of investigators reflects the workload required. ie if only one author for large cohort trial this would decrease trust; known researchers and research institutes that have robust ethical standards would increase trust"* |

| | | | |
|---|---|---|---|
| | Involvement of multidisciplinary team (including interest-holders) and collaborations | 27 | Refers to the involvement of multidisciplinary team and collaborations.<br><br>"*Authors are qualified in relevant fields (Increased trust); in addition to qualifications in study methods (eg, epidemiology, biostatistics), the research team should include subject matter experts (eg, in clinical medicine, pharmacology, environmental sampling, or other disciplines involved in collecting relevant data and applying study findings.*"<br><br>*"The involvement of relevant stakeholders (e.g., patients, clinicians, policymakers) in the design and conduct of the study can increase its relevance and trustworthiness."*<br><br>*"The group of researchers: Inclusion of biostatisticians or epidemiologists would result in higher trust than study solely done by clinicians or researchers who have previously done little observational research."*<br><br>*"Single center vs multi-center collaborative work; would trust the latter more"*<br><br>*"Increase trust when conducted in multi-country and multi-institutional"*<br><br>*"DEI [diversity equity inclusion]: increase trust"* |
| Country | Good country reputation | 6 | Refers to the research reputation of the country.<br><br>*"Authors from countries associated with poor quality research (decrease trust)"*<br><br>*"Some countries tend to be more careful than researchers from other countries"* |
| | Appropriate regulations and standards | 3 | Refers to the country's regulations and standards.<br><br>*"Region of First Author or Study Location: Concerns may arise when studies are conducted in regions where regulatory oversight is weaker."* |

| | | | |
|---|---|---|---|
| Journal | Good journal reputation (metrics, properties) | 52 | Refers to the journal reputation. Respondents noted the journal metrics (e.g., impact factor, quartile, etc) and properties such as being indexed.<br><br>*“The journal/publisher of the study and its specifications (i.e. impact factor, quartile, etc), reputable journals that is indexed and by reputable publishing facility would increase my trust and vice versa.”*<br><br>*“When the studies are published in non-reputed or predatory journal, I'm less willing to trust them.”* |
| | Appropriate peer review process | 35 | Refers to appropriate peer review process. Respondents noted the rigor of the peer review process and having open peer review.<br><br>*“My judgment of the rigor of peer review of the publishing journal. (Less rigorous would decrease trust). This is not just a matter of impact factor or "tier" of the journal, but reflects the resources available to the journal and my knowledge of and judgment of the skill, integrity, and dedication of the editor-in-chief.”*<br><br>*“Increase trust: known journal thus have some sense and trust of peer review, editorial policies”*<br><br>*“The reputation of the journal: whether it has editors with a good level and track reputation, and a good network of peer reviewers.”*<br><br>*“Open peer review that appears robust increases trust”* |
| | Open practices | 2 | Refers to the open practices of the journal, regarding being open-access, and the requirements for sharing the data and codes.<br><br>*“Published in a fully open access journal. Many journals that are fully open access publish a lot of low-quality research after questionable peer review.”*<br><br>*“Reputation and quality of journal are also a significant factors, since peer review quality varies widely and journal requirements for data and code sharing also vary”* |

| | | | |
|---|---|---|---|
| **Artificial intelligence** | | | |
| | No suspicion of use of AI for the analysis | 3 | Refers to the use of artificial intelligence for the analysis.<br><br>*“Is there any AI and/or machine learning* - this would somewhat decrease my *confidence”*<br><br>*“If the write up and/or analysis of the results used AI, then it would likely* decrease my *trust in the findings of the NRSI.”*<br><br>*“I would tend to accept the clear analytic approach over the use of AI to "determine" associations purely based on statistical patterns of association.”* |
| | No suspicion of use of AI for writing | 2 | Refers to the use of artificial intelligence for the writing of the study article.<br><br>*“[would decrease trust] if odd phrases and/or out*-of-context words appear, are they typical of those found in AI-*generated content”*<br><br>*“If the write up and/or analysis of the results used AI, then it would likely* decrease my *trust in the findings of the NRSI.”* |

**Table S6.** Completeness of reporting (N=130)

| **Completeness of reporting** | **n** | **%** |
|---|---|---|
| Quality of reporting | 19 | 14.6 |
| Adequate reporting of study methodology | 8 | 6.2 |
| Transparency in reporting | 7 | 5.4 |
| Presence of declaration statement | 7 | 5.4 |
| Adequate reporting of statistical analysis | 6 | 4.6 |
| Reporting deviations from protocol | 6 | 4.6 |
| Adequate reporting of results | 6 | 4.6 |
| Description of intervention | 6 | 4.6 |
| Adequate reporting of data sources and their quality | 5 | 3.8 |
| Adequate reporting of data collection | 3 | 2.3 |
| Adequate reporting of participant recruitment (e.g., refusals, flowchart) | 3 | 2.3 |
| Disclosure of the funding source | 3 | 2.3 |
| Availability of appendices | 3 | 2.3 |
| Adequate reporting of study design | 3 | 2.3 |
| Adequate reporting of study variables | 2 | 1.5 |
| Adequate reporting of treatment assignment | 2 | 1.5 |
| Reporting of sample size calculation | 1 | 0.8 |
| Adequate reporting of missing data | 1 | 0.8 |
| Reporting of study period | 1 | 0.8 |
| Reporting of contact details of investigators | 1 | 0.8 |
| Reporting of outcomes and measurement tools | 1 | 0.8 |
| Not reported | 36 | 27.7 |

**Table S7.** Themes according to participant characteristics (N=130)

| Characteristic | | Theme | | | | | | | | | | | | | | | |
|---|---|---|---|---|---|---|---|---|---|---|---|---|---|---|---|---|---|
| | | Open science | | Research question | | Study methodology | | Data source | | Findings and interpretation | | Writing | | Oversight | | Artificial intelligence | |
| | | Yes | No | Yes | No | Yes | No | Yes | No | Yes | No | Yes | No | Yes | No | Yes | No |
| **Age** | Mean (SD) | 52.0 (14.0) | 52.2 (14.7) | 57.4 (13.3) | 50.8 (14.0) | 49.6 (14.3) | 56.0 (12.9) | 53.1 (13.9) | 51.5 (14.2) | 52.2 (14.9) | 51.7 (12.8) | 52.1 (15.5) | 52.0 (13.7) | 50.1 (14.0) | 55.7 (13.6) | 59.5 (7.7) | 51.8 (14.2) |
| **Years of experience** | >20 years | 57 (78.1) | 16 (21.9) | 15 (20.5) | 58 (79.5) | 39 (53.4) | 34 (46.6) | 25 (34.2) | 48 (65.8) | 42 (57.5) | 31 (42.5) | 17 (23.3) | 56 (76.7) | 44 (60.3) | 29 (39.7) | 3 (4.1) | 70 (95.9) |
| | ≤20 years | 49 (86.0) | 8 (14.0) | 9 (15.8) | 48 (84.2) | 42 (73.7) | 15 (26.3) | 17 (29.8) | 40 (70.2) | 33 (57.9) | 24 (42.1) | 13 (22.8) | 44 (77.2) | 43 (75.4) | 14 (24.6) | 1 (1.8) | 56 (98.2) |
| **Level of expertise** | Advanced | 59 (75.6) | 19 (24.4) | 17 (21.8) | 61 (78.2) | 46 (59.0) | 32 (41.0) | 28 (35.9) | 50 (64.1) | 46 (59.0) | 32 (41.0) | 13 (16.7) | 65 (83.3) | 48 (61.5) | 30 (38.5) | 2 (2.6) | 76 (97.4) |
| | Lower | 47 (90.4) | 5 (9.6) | 7 (13.5) | 45 (86.5) | 35 (67.3) | 17 (32.7) | 14 (26.9) | 38 (73.1) | 29 (55.8) | 23 (44.2) | 17 (32.7) | 35 (67.3) | 39 (75.0) | 13 (25.0) | 2 (3.8) | 50 (96.2) |
| **Involvement:conducting NRSIs** | Yes | 57 (73.1) | 21 (26.9) | 16 (20.5) | 62 (79.5) | 52 (66.7) | 26 (33.3) | 28 (35.9) | 50 (64.1) | 52 (66.7) | 26 (33.3) | 15 (19.2) | 63 (80.8) | 47 (60.3) | 31 (39.7) | 3 (3.8) | 75 (96.2) |
| | No | 49 (94.2) | 3 (5.8) | 8 (15.4) | 44 (84.6) | 29 (55.8) | 23 (44.2) | 14 (26.9) | 38 (73.1) | 23 (44.2) | 29 (55.8) | 15 (28.8) | 37 (71.2) | 40 (76.9) | 12 (23.1) | 1 (1.9) | 51 (98.1) |
| **Involvement: assessing NRSIs** | Yes | 86 (83.5) | 17 (16.5) | 15 (14.6) | 88 (85.4) | 64 (62.1) | 39 (37.9) | 30 (29.1) | 73 (70.9) | 57 (55.3) | 46 (44.7) | 24 (23.3) | 79 (76.7) | 72 (69.9) | 31 (30.1) | 1 (1.0) | 102 (99.0) |
| | No | 20 (74.1) | 7 (25.9) | 9 (33.3) | 18 (66.7) | 17 (63.0) | 10 (37.0) | 12 (44.4) | 15 (55.6) | 18 (66.7) | 9 (33.3) | 6 (22.2) | 21 (77.8) | 15 (55.6) | 12 (44.4) | 3 (11.1) | 24 (88.9) |
| **Involvement: development of reporting guidelines** | Yes | 31 (88.6) | 4 (11.4) | 10 (28.6) | 25 (71.4) | 17 (48.6) | 18 (51.4) | 11 (31.4) | 24 (68.6) | 21 (60.0) | 14 (40.0) | 11 (31.4) | 24 (68.6) | 22 (62.9) | 13 (37.1) | 2 (5.7) | 33 (94.3) |
| | No | 75 (78.9) | 20 (21.1) | 14 (14.7) | 81 (85.3) | 64 (67.4) | 31 (32.6) | 31 (32.6) | 64 (67.4) | 54 (56.8) | 41 (43.2) | 19 (20.0) | 76 (80) | 65 (68.4) | 30 (31.6) | 2 (2.1) | 93 (97.9) |

Abbreviations: SD: standard deviation

*We report the frequencies (percentages %).
