## Additional file 2 for "Factors influencing the trustworthiness of non-randomized studies of interventions: a survey of international experts"

### **Additional file 2.** Survey questions

**Additional file 2.** Survey questions

### What factors might influence the trustworthiness of non-randomized (observational) studies of interventions?

#### Why are we doing this survey?

The prominence of non-randomized studies of the effects of interventions (NRSI), i.e., observational studies assessing the effectiveness of interventions, has increased in recent years, especially with the surge of large routinely collected data.

Tools have been developed to assess the risk of bias of these studies (e.g., ROBINS-I). However, as researchers, our trust in the results of reports of NRSI can be influenced by factors other than the risk of bias. These factors can pertain to the study itself, but also external factors such as the field where it is performed, the circumstances under which it is performed, the journal where it is published or other.

#### What is our aim?

The aim of the survey is to identify the factors that might influence the trustworthiness of non-randomized studies of interventions.

Trustworthiness is defined as the proper, justified or rational trust in the study findings, i.e. the findings of a study are trustworthy if there is good reason to think that these findings are true or at least sufficiently close to the truth and that they are based on sufficient high-quality evidence.

The protocol is registered on OSF and can be accessed **here**.

#### What will be your role?

It would be great if you could list factors that may influence your trust in non-randomized studies of interventions. Completing the survey will take approximately 10-15 minutes. Only de-identified data will be collected and analyzed, and the collected data will remain confidential and anonymous.

If you agree, you will be acknowledged in the resulting manuscript, provided you share your name and affiliation at the end of the survey.

Should you have any questions about this study at any time, kindly contact the research team.

**If you voluntarily agree to take part in the study; you can access the survey by clicking NEXT.**

**DEMOGRAPHICS**

1. Age:
2. How would you best describe your gender?
   - Man

- Woman
- I use a different term : [Type here]

3. Resident country: [Dropdown menu of all countries]
4. Highest educational qualification:
   - Master’s degree
   - Doctoral degree (PhD)
   - Doctor of Medicine (MD)
   - Other: Specify
5. Main field of expertise/current professional activity: (Select one)
   - Clinician
   - Methodologist
   - Epidemiologist
   - Editor
   - Systematic reviewer
   - Statistician
   - Other: Specify
6. Years of experience as a researcher:
   - 1-5 years
   - 6-10 years
   - 11-20 years
   - 21-40 years
   - > 40 years
7. Level of expertise in non-randomized studies of interventions (observational studies):
   - Novice
   - Beginner
   - Intermediate
   - Advanced
   - Expert
8. Involvement in non-randomized studies of interventions (observational studies) through: (Select all that apply)
   - Conducting non-randomized studies of interventions
   - Assessing non-randomized studies of interventions as part of evidence synthesis
   - Developing reporting guidelines for non-randomized studies of interventions
   - Other: Specify

### FACTORS

As researchers, we regularly read scientific publications and experience varying levels of trust in the reported results. This trust is based on our expertise in the planning, conduct, and reporting of studies. Besides possible biases, our trust in the study might be influenced by other factors. These factors could be related to the study itself but also to external factors such as the research field, the topic, where it is published (e.g., journal, platforms), who conducted the study, etc.

Overall, the trust in the reported results is based on implicit factors which, to our knowledge, have never been explored.

Let us illustrate with this example:

Two non-randomized (observational) studies assessing the effect of a pharmacologic treatment on patients' health have been critically appraised using a specific risk of bias tool (i.e., ROBINS-I) which explores risk of bias due to confounding; risk of bias in selection of participants; risk of bias in classification of interventions; risk of bias due to deviations from intended interventions; risk of bias due to missing data; risk of bias in measurement of outcomes; and risk of bias in selection of the reported result. Similar issues were identified in both studies and they were both rated as having an overall moderate risk of bias.

However, when you read the reports, you believe that one study is more trustworthy than the other. This judgment is based on factors, other than the risk of bias, that influence your trust.

With this in mind, please think about recent reports of NRSI that you have read, and identify the factors, other than the risk of bias, that might influence your trust in the results of the study.

You may list factors that pertain to the study itself (e.g., quality of the dataset), who led, conducted, analyzed or reported the study (e.g., reputation of the research centers), where it is published (e.g., predatory journals), the field where it is performed, the circumstances under which it is performed, the research practices (e.g., data sharing), the effect estimates (e.g., unexpectedly large effect estimates) or any other factors that you may consider influential.

Please list all factors in your own words and provide a brief explanation and specify whether the factor might increase or decrease your trust in non-randomized studies of interventions. We invite you to cite as many factors you think might influence your trust in the study.

Of note, some factors might be rarely reported in non-randomized studies of interventions, but should nevertheless be considered. If you have any doubt on whether the factor is covered in the risk of bias, then include it in the list of factors below.

#### Factor 1

Please indicate the factor, provide a brief explanation and specify whether the factor might increase or decrease your trust in a non-randomized (observational) study of interventions.

etc

**Factor 10**

Please indicate the factor, provide a brief explanation and specify whether the factor might increase or decrease your trust in a non-randomized (observational) study of interventions.

**Additional factors** (if needed)

Please indicate the factor(s), provide a brief explanation and specify whether the factor(s) might increase or decrease your trust in a non-randomized (observational) study of interventions.

### ACKNOWLEDGEMENT

Do you want to be acknowledged in our publication?

- Yes, I want to be acknowledged

    → Please provide your name, email address and affiliation.

- No, I do not want to be acknowledged

### THANK YOU
